# Vitamin D and Prebiotics for Intestinal Health in Cystic Fibrosis: Rationale and design for a randomized, placebo-controlled, double-blind, 2 × 2 trial of administration of prebiotics and cholecalciferol (vitamin D_3_) (Pre-D Trial) in adults with cystic fibrosis

**DOI:** 10.1101/2024.01.04.24300860

**Authors:** Alisa K. Sivapiromrat, Pichatorn Suppakitjanusant, Yanling Wang, Jose Binongo, William R. Hunt, Andrew Gewirtz, Jessica A. Alvarez, Chengcheng Hu, Samuel Weinstein, Ishaan Jathal, Thomas R. Ziegler, Vin Tangpricha

## Abstract

Individuals with cystic fibrosis (CF) have dysfunctional intestinal microbiota and increased gastrointestinal (GI) inflammation also known as GI dysbiosis. It is hypothesized that administration of high-dose cholecalciferol (vitamin D_3_) together with a prebiotic (inulin) will be effective, and possibly additive or synergistic, in reducing CF-related GI dysbiosis and improving intestinal functions. Thus, a 2 × 2 factorial design, placebo-controlled, double-blind, clinical trial was proposed to test this hypothesis. Forty adult participants with CF will be block-randomized into one of four groups: 1) high-dose oral vitamin D_3_ (50,000 IU weekly) plus oral prebiotic placebo daily; 2) oral prebiotic (12 g inulin daily) plus oral placebo vitamin D_3_ weekly; 3) combined oral vitamin D_3_ weekly and oral prebiotic inulin daily; and 4) oral vitamin D_3_ placebo weekly and oral prebiotic placebo. The primary endpoints will include 12-week changes in the reduced relative abundance of gammaproteobacteria, and gut microbiota richness and diversity before and after the intervention. This clinical study will examine whether vitamin D_3_ with or without prebiotics will improve intestinal health and reduce GI dysbiosis, which in turn, should improve health outcomes and quality of life of patients with CF.

## 1. Introduction and Background

Cystic fibrosis (CF) is the most common life-shortening genetic disease among the Caucasian population in the United States [1]. CF is caused by a mutation in the cystic fibrosis transmembrane conductance regulator (CFTR) gene, which results in the disruption of water and chloride ion transport on epithelial surfaces, such as the lung, liver, pancreas, and gastrointestinal tract [2]. As a result, patients with CF have altered gastrointestinal (GI) microbiota, consisting of reduced diversity of their GI microbiota profile (dysbiosis) and increased pro-inflammatory microbiota in comparison to people without CF [3]. Further, the CFTR mutation type influences the severity of GI dysbiosis [7, 10, 11].

Additional factors that may contribute to dysbiosis include recurrent exposure to antibiotics, persistent infection and inflammation, and the traditional high-fat and high-calorie CF diet [4, 5, 33, 35]. GI microbiota dysbiosis is further aggravated by insufficient production of pancreatic enzymes, resulting in fat and protein malabsorption [6]. Further, GI dysbiosis is associated with small bowel bacterial overgrowth, altered bowel motility, pancreatic insufficiency, and inflammation of the bowel [6, 8, 9]. The culmination of these factors contributes to the aberrant intestinal microbiota. GI dysbiosis is associated with significant CF morbidity, including lowered pulmonary function, heightened need for IV antibiotics, and impaired growth in children [8, 12, 13]. Therefore, improving GI microbiota dysbiosis may be beneficial to improving the quality of life of CF patients.

Prebiotics are non-digestible oligosaccharides derived from a variety of plants and may modulate and improve GI microbiota diversity and inflammation [9, 14]. Inulin is broken down by GI microbiota and is derived from chicory root; it is the water-soluble prebiotic chosen for this clinical trial due to its beneficial role in GI health [15, 16]. Although previous studies have demonstrated that prebiotics may have efficacy in promoting beneficial GI microbiota and improving respiratory infections in adults and children without CF, no studies have been published regarding the administration of prebiotics in adults with CF [27, 28, 29, 30, 31].

GI dysbiosis and inflammation are associated with vitamin D deficiency, which is commonly seen in patients with CF [21]. Previous randomized clinical trials (RCTs) showed that vitamin D administration has been linked to increased GI health-promoting bacteria, decreased GI pathogenic bacteria, and improved airway microbiota [19, 20, 21]. Administration of vitamin D is associated with increased GI microbiota diversity and composition [17, 18, 19, 20, 21].

Thus, it is hypothesized that a high weekly dose of oral vitamin D_3_ and oral prebiotic inulin will effectively decrease GI dysbiosis in participants with CF after 12 weeks, including possible synergistic effects of the vitamin D_3_ and prebiotic combination. This will be a 2 × 2, placebo-controlled, randomized, double-blind, clinical study to examine oral vitamin D_3_ and prebiotic administration in adults with CF and the changes in GI microbiota richness and diversity before and after study intervention.

## 2. Materials and methods

### 2.1. Overview of study design

This study was designed as a pilot and feasibility study. The design of the study was a 2 × 2, a double-blind, randomized, placebo-controlled clinical trial to define the role of vitamin D_3_ and prebiotic (inulin) supplementation on GI dysbiosis in patients with CF (Figure 1). Adults with CF were randomized to 1) oral vitamin D_3_ weekly plus oral prebiotic placebo daily; 2) oral prebiotic daily plus oral placebo vitamin D_3_ weekly; 3) combined oral vitamin D_3_ weekly and oral prebiotic daily; and 4) oral placebo vitamin D_3_ weekly and oral placebo prebiotic daily. The prebiotic inulin and placebo inulin were dissolved in a liquid beverage of the participant’s choice and ingested daily. The participants were allowed to continue taking vitamin D if they were already on maintenance vitamin D therapy as long as the supplemental vitamin D_3_ dose remained no greater than 2000 IU daily. Subjects were followed for 12 weeks for clinical trial endpoints. Questionnaires on vitamin D intake and medical history were administered at baseline and the final visit. Biweekly follow-up phone calls were made to assess for potential adverse effects. Relevant clinical outcome study endpoints were extracted from the electronic medical record.

**Figure 1.**
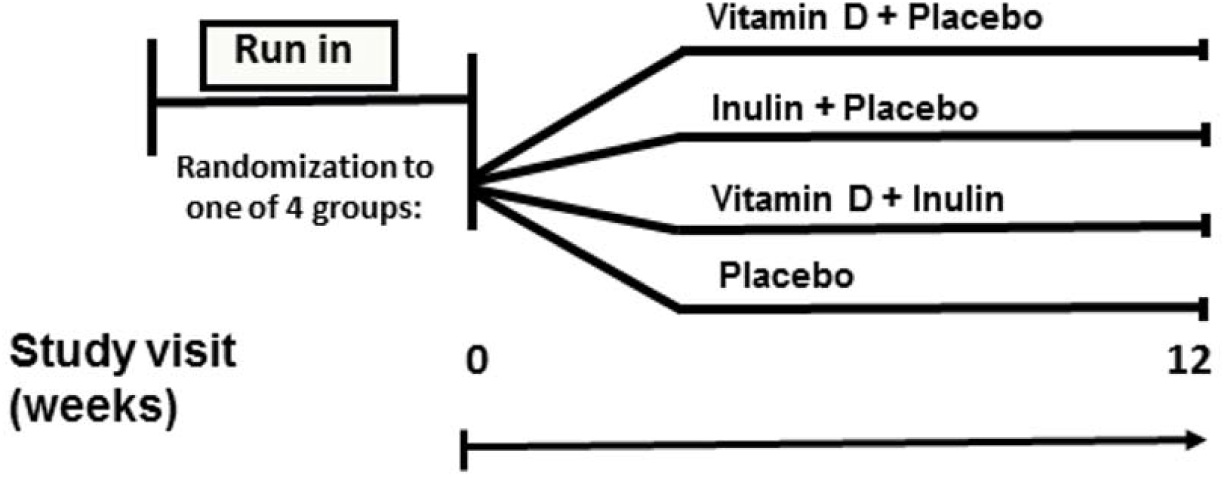
Overview study design of the Vitamin D and Prebiotics for Intestinal Health in Cystic Fibrosis (Pre-D) clinical trial

### 2.2. Hypothesis and aims

We hypothesize that the administration of high-dose vitamin D_3_ and prebiotic will decrease the relative abundance of gammaproteobacteria and reduce GI dysbiosis in adults with CF, including cumulative or synergistic effects of the vitamin D_3_ and prebiotic combination. Sputum will be collected as an exploratory endpoint to examine the impact of the intervention on airway microbiota.

### 2.3. Sponsors

The primary sponsor of “Vitamin D and Prebiotics for Intestinal Health in Cystic Fibrosis” is the CF Foundation and the National Institutes of Health (NIH).

### 2.4. Setting and clinical sites

This clinical study was conducted at the Emory Adult Cystic Fibrosis Center, Atlanta, GA, a United States-based CF Foundation accredited Care Center, and member of the CF Therapeutics Development Network (TDN). All study participants received clinical care for CF at this center.

### 2.5. Intervention arms: Vitamin D_3_ supplement, prebiotic, both vitamin D_3_ and prebiotic supplement, and placebo

Patients were randomized into a 2 × 2 factorial design, placebo-controlled clinical trial with 4 interventional branches: 1) oral vitamin D_3_ (50,000 IU) weekly and oral corn-derived maltodextrin daily (12 g as the oral prebiotic placebo); 2) oral placebo vitamin D_3_ weekly and oral chicory-derived inulin (12 g) prebiotic daily ; 3) oral vitamin D_3_ weekly and oral inulin prebiotic daily; and 4) oral placebo vitamin D_3_ weekly and oral placebo prebiotic daily. The vitamin D_3_ and matching identical placebo capsules were obtained from BioTech Pharmacal Inc. (Fayetteville, AR), and the inulin was obtained from Now Foods (Bloomingdale, IL). An independent laboratory (ARL Bio Pharma, Oklahoma City, OK) analyzed the vitamin D_3_ capsules to confirm the dosage and ensure the product remained stable throughout the duration of the study. The Investigational Drug Service at Emory University Hospital stored and dispensed the study medication at baseline as 1 pill weekly (50,000 IU of vitamin D_3_ or placebo) and 1 powder packet daily (12 g of inulin or placebo) for the duration of the study (12 weeks).

### 2.6 Trial eligibility

The following criteria were used to determine eligibility for participation in the “Vitamin D and Prebiotics for Intestinal Health in Cystic Fibrosis.” Inclusion criteria: 1) male and female patients (18 years old or older) with confirmed CF by genetic mutation and/or sweat chloride testing, 2) not currently on oral or systemic antibiotics for pulmonary exacerbation or any other reason, including chronic use of oral azithromycin (or macrolide therapy) (minimum of four weeks prior to starting study) – chronically inhaled antibiotics were allowed as long as the participants had not started the inhaled antibiotic less than four weeks from study enrollment, 3) use of CFTR modulator therapy is allowed. Exclusion Criteria: 1) active GI disease, abdominal pain and/or diarrhea, 2) chronic kidney disease worse than stage 3 (eGFR < ml/min per 1.73 m^2^), 3) any vitamin D supplement or vitamin D analogue use greater than 2,000 IU (patients who are taking more than 2,000 IU of vitamin D must agree to stop the vitamin D for 6 weeks and take less than 2,000 IU of vitamin D during the study), 4) use of immunosuppressants or history of organ transplantation, 5) current use of prebiotics or probiotics.

### 2.7 Recruitment of the study population

Potential subjects were screened for eligibility based on inclusion and exclusion criteria by utilizing electronic medical records before enrollment (Table 2A). Subjects were interviewed during their visit to the clinic for further screening and approached for informed consent if they were eligible for the study. Study participants were either enrolled in the study or reported as screen failures (Figure 2).

**Table 2. (A).** Reasons for exclusion and ineligibility for all patients screened (n = 214).

| Reasons for exclusion and ineligibility | Excluded (n = 214) |
| --- | --- |
| - Declined to participate | 81 |
| - Did not meet inclusion criteria | 41 |
| - Not diagnosed with cystic fibrosis | 9 |
| - Taking antibiotics for pulmonary exacerbation or any other reason | 32 |
| - Met exclusion criteria | 65 |
| - Vitamin D<sub>3</sub> intake > 2,000IU | 41 |
| - Taking probiotics or prebiotics | 5 |
| - Enrolled in another conflicting or interventional study | 7 |
| - Too sick to participate in study | 10 |
| - On cytotoxic or immunosuppressive drugs | 2 |
| - Did not return first specimen samples | 16 |
| - Other/unknown | 11 |

**Figure 2.**
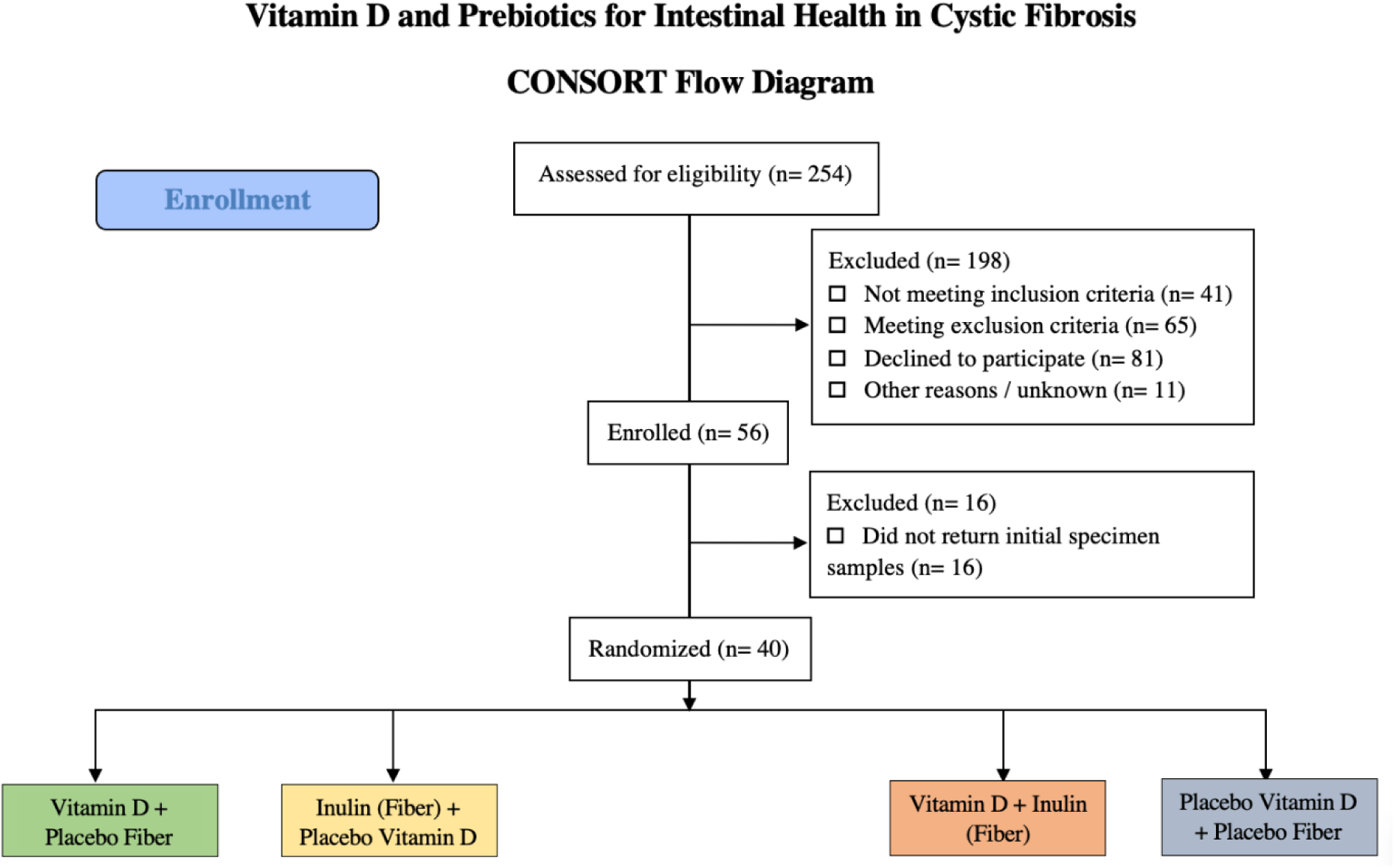
CONSORT diagram of randomized Vitamin D_3_ and Prebiotics for Intestinal Health in Cystic Fibrosis (Pre-D) clinical trial.

#### 2.7.1. Enrollment

Before participating in the study procedures, all study participants provided written informed consent. Forty subjects were randomized into the study and given a demographics questionnaire (Table 2B).

**Table 2. (B).** Demographics for subjects randomized in the 12-week Pre-D clinical trial (n = 40).

| Demographics | Patients (n = 40) |
| --- | --- |
| Age | 31.5 ± 11.0 |
| BMI (kg/m<sup>2</sup>) | 24.4 ± 4.1 |
| Height (cm) | 167.0 ± 10.0 |
| Weight (kg) | 68.3 ± 14.1 |
| Sex (%) |  |
| - Male | 57.5% |
| - Female | 42.5% |
| Race (%) |  |
| - Caucasian | 90% |
| - African American | 7.5% |
| - Native Hawaiian/Pacific Islander | 2.5% |

#### 2.7.2. Randomization

Forty CF adult subjects were block-randomized to one of the four groups (10 subjects per interventional arm). Randomization was performed by a biostatistician not involved in subject recruitment. Investigational Drug Service research pharmacists dispensed the study drugs with new labels to keep the study blinded to all study investigators, staff, and subjects.

### 2.8. Specimen collection and assays

#### 2.8.1. Stool and sputum collection and processing

Subjects provided stool and sputum samples via return mail at baseline and endpoint (12 weeks). Stool and sputum were collected prior to starting medication and collected again after completion of medication. Subjects were provided two stool collection kits (OMR-200, OMNIgene Gut sample collection kit, Ottawa, Ontario, Canada) and two sputum collection kits (standard 50 mL conical tube) on the baseline visit day to mail directly to the lab within 48 hours after each collection. The OMR-200 kit provides microbiome stability at room temperature within 60 days of collection (reference https://www.dnagenotek.com/row/pdf/PD-BR-00181.pdf). Non-induced expectorated sputum was produced and collected by subjects upon coughing. Analysis was done if both baseline and endpoint samples were returned. If none or only one of the baseline or endpoint samples were returned, samples were excluded from the analysis.

##### 2.8.1.1 Stool and sputum fluid-based assays

DNA will be extracted and purified from the frozen stool and sputum samples using DNeasy 96 PowerSoil Pro QIAcube HT kit, supplemented with PowerBead Pro Plates (Qiagen). The sequencing library will be made following Illumina 16S Metagenomic Sequencing Library Preparation (https://support.illumina.com/documents/documentation/chemistry_documentation/16s/16s-metagenomic-library-prep-guide-15044223-b.pdf). Briefly, V3-V4 region of the 16s rRNA genes of each sample will be amplified. Amplicons will be verified on an agarose gel and purified using AMPure XP magnetic beads (Beckman). A second PCR will be performed to attach dual indices and Illumina sequencing adapters using Nextera XT Index Kit (Illumina). Amplicons will be verified and purified again as stated above. The final purified amplicons will be quantified using Quant-iT™ PicoGreen™ dsDNA Assay Kits (ThermoFisher). An equal molar of amplicons of each sample will then be combined. The combined product will be diluted and spiked with 10% PhiX control (Illumina) and sequenced by Illumina MiSeq sequencer (2 × 250bp). Sequence data will be analyzed using Qiime 2, following the protocols of the Qiime2 Moving Pictures tutorial (https://docs.qiime2.org/2022.2/tutorials/moving-pictures/). Briefly, sequences will be demultiplexed and quality filtered with the DADA2 plugin. Alpha and beta diversity will be analyzed using the phylogenetic tree built by the FastTree program in Qiime2. Taxonomy will be assigned using Greengenes database 13_8_99. Linear discriminate analysis effect size (LEfSe) will be done to determine bacterial members that explain differences between groups.

### 2.9. Follow-up and study endpoint determination

#### 2.9.1 Primary endpoint determination (improved GI and lung microbiota composition and diversity)

At the conclusion of the 12-week RCT, changes in GI and lung microbiota were determined using 16S rRNA gene sequencing and microbiome-dependent metabolites pathways in stool and sputum using high-resolution metabolomics analysis. The primary endpoint determines if vitamin D_3_ and a prebiotic improved species richness alpha diversity (Shannon Index) and the reduced relative abundance of gammaproteobacteria, improving the health of the subjects.

#### 2.9.2 Secondary endpoint determination (changes in GI and lung microbiota composition and diversity, and clinical pilot study tolerability)

Secondary endpoints include changes in GI diversity and richness from any interventional arm in a direction of no health consequence. Future investigations may explore markers of glucose metabolism concerning the most common CF comorbidity: cystic fibrosis-related diabetes (CFRD) [22]. Another secondary endpoint was to establish the tolerability of this pilot study to determine the feasibility of similar larger clinical studies in the future.

### 2.10. Adherence and tolerability assessment

Patient adherence to the study intervention regimen was assessed via bi-weekly telephone calls to verify the pills and packets had been taken as prescribed, determine missed dosages, and remind patients of study drug adherence. At the end of the study, a telephone call was made to inquire about the number of remaining pills and the number of packets.

Subjects were asked “How acceptable was the study medication on a scale from 1 to 5?” to assess the tolerability of the study medication after study completion. Subjects scored tolerability on a scale ranging from 1 to 5: 1 = not acceptable; 2 = moderately not acceptable; 3 = somewhat acceptable; 4 = moderately acceptable; 5 = highly acceptable.

### 2.11. Data management

All clinical trial data was recorded on paper case report forms and transferred to the Research Electronic Data Capture (REDCap), a secure electronic database for clinical and translational research. The clinical trial data was entered throughout the study. The REDCap database was reviewed to ensure the data quality is sustained. The study PI completed a final review of case report forms and REDCap database entry to confirm all clinical trial data matches the source paper case report documentation. A not for cause Emory IRB audit confirmed the integrity of the study procedures and data management in October 2022.

### 2.12. Sample size determination and analysis plan

#### 2.12.1. Analysis Plan

The sample size (n = 40) of this pilot study was based on the pragmatics of recruitment, patient flow, and budgetary constraints. Preliminary data from the vitamin D + placebo arm suggested a standard deviation of 1.2 of the Shannon diversity index. Vitamin D arms (vitamin D and prebiotic placebo, and vitamin D and inulin prebiotic arms) will have 29% power to detect a between-group difference of 0.5, and 56% power to detect a difference of 0.75 in the diversity index using a significance level of 0.05. Power calculation of the prebiotic arms (prebiotic inulin and placebo vitamin D, and prebiotic inulin and vitamin D arms) was not available. However, if the vitamin D and pre-biotic combination are shown to have a synergistic effect, there may be greater power to detect a between-group difference of 0.5 (or 0.75). As a pilot study, the primary goal is not to evaluate the efficacy of the proposed interventions but to examine the feasibility of conducting this study on a larger scale. Data will provide a first impression of the variability and effect sizes of the Shannon diversity index in the patient population of interest in each of the four treatment arms, inform power calculations for secondary outcomes, assist in the sample size and power calculations of the subsequent larger study while helping identify future modifications. Between-group differences will be determined with an understanding that low power is characteristic of pilot studies. Patient demographics were summarized using mean ± SD (or median ± interquartile range, as needed) for continuous variables and frequency for categorical variables, and diversity index statistics will be summarized at baseline and endpoint for each arm. The feasibility of recruitment, retention, assessment, and implementation of the proposed interventions will be evaluated to identify any potential flaws in the study design and potential challenges in the subsequent large-scale study. Feasibility measures will focus on metrics of recruitment, retention, acceptability, and completion as adopted from the NIH (National Institutes of Health). The target metric of success is compared to our previous single intervention study with vitamin D.

### 2.13. Ethics and data safety

#### 2.13.1. Human subjects (IRB) – consent and confidentiality

The clinical study was approved by the Emory University Institutional Review Board (IRB). The study was registered at clinicaltrials.gov (<u>NCT04118010</u>). As required by Emory University IRB regulations, all study investigators and staff completed proper clinical trials and research training. Case report forms for clinical trial data were stored in locked storage spaces. Data was entered into REDCap in real-time throughout the clinical study.

Potential subjects were patients seeking care as outpatients at the Emory University Adult Cystic Fibrosis Clinic. Patients were screened and enrolled through an informed consent process. There were two consent forms: 1) Consent to participate as a research subject; 2) HIPAA compliance measures are described to participants by trained study investigators. Subjects were encouraged to actively discuss their understanding and to inquire about any questions. All participants were given signed consent form copies and contact information of the study team in case of adverse events or intent to withdraw participation in the study.

## 3. Discussion

The purpose of this pilot study will be to determine the role of vitamin D_3_ combined with a prebiotic in outpatient adults with CF. The study will examine GI and lung microbiota richness and diversity changes between subjects administered vitamin D_3_, prebiotic, combined vitamin D_3_ and prebiotic, and placebo vitamin D_3_ and placebo prebiotic.

The severity of GI dysbiosis is associated with the CFTR mutation type [7, 10, 11]. In a study of adolescents with CF, the severity of the CFTR mutation (presence of F508del) was associated with more abundant pathogenic bacteria, less abundant beneficial bacteria, and less microbial diversity, suggesting an inherent dysbiosis caused by the CFTR defect [10]. Rodent models of CF mirrored GI dysbiosis in patients with CF; those with CFTR mutations have less bacterial diversity and richness compared to their wild-type (WT) counterparts [11, 34].

Up to 90% of patients with CF have vitamin D insufficiency (serum 25-hydroxyvitamin D (25(OH)D < 30ng/mL) [23]. As such, the Cystic Fibrosis Foundation recommends annual measurement of serum 25(OH)D concentration [37, 38]. Vitamin D is associated with better skeletal health, lung function, and decreased risk of pulmonary exacerbations, cystic fibrosis-related diabetes, and infections [23, 37, 39, 40, 41]. Bolus vitamin D dosages are often required to increase serum 25(OH)D concentration in patients with CF, as patient adherence to other treatment options is lower [24, 25, 26]. Given updated CF Foundation recommendations, the vitamin D status of patients with CF has improved; however, how vitamin D therapy affects clinical outcomes in CF, such as GI dysbiosis and intestinal functions, is still poorly understood.

Vitamin D deficiency has been associated with GI dysbiosis. In rodent models of CF, vitamin D deficient mice or vitamin D receptor KO mice have increased intestinal dysbiosis and GI inflammation [21]. Kanhere et al found that participants with CF who received vitamin D demonstrated fewer pathogenic bacteria (gammaproteobacteria) and increased traditionally less pathogenic bacteria [19]. Further, participants with CF also had improved GI and airway microbiota after weekly vitamin D administration [19]. Another study in hospitalized adults with CF found that GI microbial metabolism was altered following a bolus dose of vitamin D [20]. Vitamin D administration has been linked to increased health-promoting species and altered microbiota composition and diversity [19, 20, 21]. Whether the addition of a prebiotic improves GI or lung microbiota is a focus of this study.

Several RCTs demonstrated prebiotics alter GI microbiota in adults and children without CF [27, 28, 29]. Prebiotics potentially has a better safety profile than probiotics since they do not contain bacteria, and thus, are not associated with bacteremia [36]. Other RCTs have demonstrated that changes in GI microbiota were associated with improved lung health [30, 31]. Ranucci et al reported that in a 24-month RCT, infants without CF given a prebiotic oligosaccharide-enriched formula were protected against respiratory infections and had an increased abundance of Bifidobacterium, a genus associated with protection against respiratory infections [30]. Li et al reported fewer episodes of respiratory infections in children when using prebiotic formula compared to a standard formula [31]. Despite previous RCTs demonstrating the efficacy of prebiotics in promoting beneficial GI microbiota and improving respiratory infections in children without CF, no clinical studies have been published using a combination of prebiotics and vitamin D in adults with CF. Vitamin D_3_ and Prebiotics for Intestinal Health in Cystic Fibrosis is the first clinical study examining this.

Forty subjects were randomized into one of the four study groups. The mean age of subjects was 31.5 ± 11.0 years with an even distribution of males (57.5%) and females (42.5%) participating in the study. Subjects were predominantly Caucasian (90%), as CF is far less common among other racial groups [32]. Final collection and analysis of the stool and sputum microbiome is currently underway.

If the combination of vitamin D_3_ and prebiotics is demonstrated to have synergistic or additive effects superior to that of the placebo or each intervention, this will provide the rationale to study this combination in a larger multi-center study or to study other combinations of pre or probiotics with vitamin D_3_. Furthermore, if the study results are positive, further investigation into the mechanism by which vitamin D_3_ and/or prebiotics improve gut and airway microbiomes will need to be explored. A limitation of this study is that blood samples were not drawn during the study due to accessibility issues caused by the COVID-19 pandemic. Therefore, data on inflammatory and gut function tests will not be available. Based on past studies using this vitamin D_3_ regimen, it is accepted that vitamin D concentrations improved with the vitamin D_3_ intervention.

Past studies have shown promising outcomes in patients with CF after vitamin D_3_ supplementation. CFRD and CF bone disease, common co-morbidities in CF, have been associated with vitamin D insufficiency and deficiency. The overall results from the Vitamin D_3_ and Prebiotics for Intestinal Health in Cystic Fibrosis (Pre-D) clinical study will inform as to whether vitamin D_3_ and prebiotics have synergistic effects on intestinal function, and GI and lung microbiota diversity and composition.

## Data Availability

All data produced in the present study are available upon reasonable request to the authors.

## Acknowledgments

The study investigators would like to acknowledge the CF care team at the Emory Adult Cystic Fibrosis and the research study coordinators for making this study feasible. This study was supported by CFF Awards TANGPR19A0 and CC002-AD and NIH awards P30DK125013 and 3UL1TR002378-05S2.

